# SSRI prescription during acute COVID-19 and risk of Long COVID symptoms and conditions among patients with depression

**DOI:** 10.64898/2026.07.06.26357401

**Authors:** Zachary Butzin-Dozier, Yunwen Ji, Lin-Chiun Wang, Manav Kumar, A. Jerrod Anzalone, Eric Hurwitz, Ariana Budhihartanto, Rena C. Patel, Alan Hubbard, Jodi Halpern, the National Clinical Cohort Collaborative (N3C) Consortium

## Abstract

**Background:** Long COVID is a syndrome characterized by symptoms and conditions across all biological systems. This breadth of Long COVID phenotypes impedes efforts to identify the mechanistic pathways of Long COVID. Low serotonin may play a role in long-term sequelae of COVID-19, and selective serotonin reuptake inhibitors (SSRIs) may prevent these sequelae. Evaluation of the relationship between SSRIs and distinct categories of symptoms and conditions associated with Long COVID can highlight the mechanistic pathways that drive these relationships.

**Methods:** We evaluated electronic health record data from a retrospective cohort of patients in the National Clinical Cohort Collaborative with comorbid depression and COVID-19 between October 2021 and February 2024. We estimated the relationship between SSRI prescription (versus no SSRI prescription) during acute COVID-19 and the one-year cumulative incidence of Long COVID-related conditions and symptoms across 14 human phenotype ontology categories. We applied Super Learner and targeted maximum likelihood estimation to estimate risk ratios while adjusting for confounders of interest and correcting for false discoveries from repeated testing.

**Results:** We evaluated EHR data from 542,938 patients. We found that patients who were prescribed SSRIs during COVID-19 had a significantly lower risk of symptoms and conditions related to gastrointestinal factors (adjusted risk ratio (aRR) 0.95, 95% CI 0.92, 0.97), general health (aRR 0.91, 95% CI 0.88, 0.95), headaches (aRR 0.96, 95% CI 0.92, 0.99) and skin (aRR 0.92, 95% CI 0.87, 0.98).

**Discussion:** We found that the prescription of SSRIs during acute COVID-19 was associated with a significantly lower risk of post-COVID sequelae related to gastrointestinal, headache-related, skin-related, and general symptoms and conditions, compared with no SSRI prescription. These findings highlight the role of serotonin in Long COVID and specific sequelae that may be reduced by SSRIs.

## INTRODUCTION

Patients infected with SARS-CoV-2 experience a wide range of symptoms that typically subside within several weeks, but a subgroup of patients experiences long-term symptoms after the acute infection has resolved. Given the global prevalence of COVID-19, now that SARS-CoV-2 has become endemic, it is crucial to understand the pathways that drive long-term consequences of infection and how to interrupt these outcomes.

A recent study found that low serotonin may be a key driver of many post-COVID-19 symptoms.^1^ This theory highlights the biological mechanism where (1) SARS-CoV-2 infection elicits the release of virus-induced interferons, (2) these interferons prevent the uptake of tryptophan, which is a precursor to serotonin (i.e., interferons prevent the production of serotonin), (3) low serotonin impairs vagal functioning, neurologic functioning, and coagulation.^1,2^

Observational studies have found that use of SSRIs during acute COVID-19 is associated with a lower subsequent risk of Long COVID.^1–3^ On the other hand, the COVID-OUT randomized trial did not find a significant relationship between COVID-19 outpatient assignment to an SSRI and subsequent risk of Long COVID.^4^ Given the breadth of post-COVID phenotypes and the multiple hypothesized biological mechanisms that drive them, considerable effort has been devoted to identifying unique clusters of post-COVID phenotypes.^5,6^ Understanding the relationships between serotonin and unique post-COVID symptoms may provide important information on the clusters of symptoms that may be treated with SSRIs and the clusters of patients who have Long COVID driven by low serotonin.

In this study, we sought to evaluate the relationship between SSRI prescription during acute COVID-19 and the subsequent risk of post-COVID symptoms among a sample of electronic health records from patients with depression in the National Clinical Cohort Collaborative (N3C).

## METHODS

### Sample

We analyzed data from electronic health records (EHR) from patients in the National Clinical Cohort Collaborative (N3C), which is a large, national open source of health data that contains information for more than 21 million patients.^7,8^ Our sample included patients diagnosed with acute COVID-19 between October 1, 2021 and February 1, 2024 with comorbid depression. N3C defines COVID-19 positivity as (A) at least one laboratory diagnostic positive result or (B) a provider diagnosis (ICD-10-CM U07.1). We defined the index date for COVID-19 as the earliest of the two events.^9^

### Exposure

Our exposure of interest was prescription of selective serotonin reuptake inhibitors (SSRI) during acute COVID-19. To be defined as an SSRI recipient, the patient must have been prescribed an SSRI at least 30 days before acute COVID-19 and did the prescription did not stop before COVID-19. SSRIs include fluoxetine, sertraline, paroxetine, citalopram, escitalopram, fluvoxamine, and vilazodone.

### Outcomes

Our outcomes of interest were 211 conditions and symptoms associated with Long COVID (hereafter referred to as “symptoms”) between 1 and 12 months after acute COVID-19. We selected this time period at-risk in accordance with Centers for Disease Control and Prevention guidelines.^10^ These 211 symptoms were drawn from previous studies that have identified and validated their use as metrics of Long COVID (post-acute sequelae of COVID-19)^11^ from the Human Phenotype Ontology (HPO) (for full list of Long COVID symptoms and conditions, see Supplemental Material 1).^6,12–14^ We summarized these symptoms across 14 symptom categories (where each category is a binary indicator of whether a patient had any symptom in that category) and as a single binary indicator (whether the patient had any post-COVID symptom). We excluded 6 symptom categories (and 76 symptoms) associated with depression and SSRI use, as they would be more frequently evaluated in patients with depression who were prescribed SSRIs compared to patients not prescribed SSRIs (i.e., measurement bias). We excluded neuropsychiatric symptom categories related to emotion and mood, behavioral, sleep, cognitive dysfunction, memory, and findings.^15^

### Covariates

Our analysis adjusted for the following confounders at baseline (acute COVID-19): healthcare utilization rate (healthcare interactions per month before SARS-CoV-2 infection), sex, age at acute SARS-CoV-2 infection, race/ethnicity, region of residence, body mass index (BMI), tobacco smoking status, obesity, diabetes, chronic lung disease, heart failure, hypertension, use of systemic corticosteroids, depression severity, anxiety, antipsychotic medication use, benzodiazepine medication use, whether the patient was immunocompromised, the number of COVID-19 vaccination doses before infection, date of COVID-19, and Charlson Comorbidity Index.^16^ We adjusted for the following county-level socioeconomic variables: the percentage of the county below the poverty line and the county’s social deprivation index score. We considered monitoring as a source of informative right censoring, where we considered a patient as informatively censored if they had fewer than 2 healthcare interactions or died within the 12 months following acute COVID-19, and evaluated our causal parameter under a scenario of universal monitoring (i.e., no censoring) consistent with previous studie.^2,17–19^

### Analysis

We applied Super Learner and Targeted Maximum Likelihood Estimation, which are effective for this analytic setting that relies on electronic health records with a high proportion of missingness.^18–21^ We used Super Learner, an ensemble machine learning algorithm, to maximize the prediction of the outcome, post-COVID symptoms, as well as the treatment and censoring mechanisms.^17,22,23^ We included the learners: generalized linear models (“SL.glm”), GLM net (“SL.glmnet”), and XGBoost (“SL.xgboost”).^23^ Next, we applied targeted maximum likelihood estimation to estimate the risk ratio comparing the risk of a given symptom category in patients prescribed an SSRI during COVID-19 to that in patients not prescribed an SSRI during COVID-19.^18–21^ Targeted maximum likelihood estimation is a doubly-robust estimator, meaning that it provides valid estimates as long as either the outcome regression or treatment mechanism is estimated consistently.^18–21^ During the one-year follow-up period, we considered mortality and lack of healthcare utilization as sources of informative censoring.^17^ We corrected our estimates for multiple testing using the Benjamini-Hochberg correction.

## RESULTS

**Table 1.** Characteristics of patients with depression who were prescribed serotonin reuptake inhibitors during acute COVID-19.

| Characteristic | Value | SSRIs: Count<br>(Proportion) | No SSRIs: Count<br>(Proportion) | Total Count<br>(Proportion) |
| --- | --- | --- | --- | --- |
| Total |  | 134595 (0.25) | 408343 (0.75) | 542938 (1) |
| Sex | Female | 100132 (0.74) | 290954 (0.71) | 391086 (0.72) |
| Measurements | Age: mean (SD) | 51.7 (19.4) | 49.77 (19.6) | 50.25 (19.57) |
|  | BMI: mean (SD) | 34.17 (9.62) | 33.39 (9.51) | 33.59 (9.54) |
| Ethnicity | White Non-Hispanic | 101196 (0.75) | 289604 (0.71) | 390800 (0.72) |
|  | Black or African American Non-Hispanic | 15552 (0.12) | 49251 (0.12) | 64803 (0.12) |
|  | Hispanic or Latino Any Race | 9715 (0.07) | 36083 (0.09) | 45798 (0.08) |
|  | Unknown | 4024 (0.03) | 14738 (0.04) | 18762 (0.03) |
|  | Other Non-Hispanic | 781 (0.01) | 6780 (0.02) | 7561 (0.01) |
|  | American Indian Non-Hispanic | 627 (0) | 2361 (0.01) | 2988 (0.01) |
|  | Asian or Pacific Islander Non-Hispanic | 2700 (0.02) | 9526 (0.02) | 12226 (0.02) |
| Medical Conditions | Tobacco Smoker | 33202 (0.25) | 86343 (0.21) | 119545 (0.22) |
|  | Obese | 84985 (0.63) | 231129 (0.57) | 316114 (0.58) |
|  | Diabetes Uncomplicated | 36142 (0.27) | 94674 (0.23) | 130816 (0.24) |
|  | Diabetes Complicated | 26063 (0.19) | 67271 (0.16) | 93334 (0.17) |
|  | Chronic Lung Disease | 47932 (0.36) | 129312 (0.32) | 177244 (0.33) |
|  | Heart Failure | 19746 (0.15) | 45778 (0.11) | 65524 (0.12) |
|  | Hypertension | 71805 (0.53) | 193674 (0.47) | 265479 (0.49) |
|  | Anxiety | 103512 (0.77) | 259908 (0.64) | 363420 (0.67) |
|  | Immunocompromised | 20635 (0.15) | 53399 (0.13) | 74034 (0.14) |
|  | Charlson Comorbidity Index Score: mean (SD) | 2.36 (2.78) | 1.96 (2.51) | 2.06 (2.59) |
| Depression | Mild Depression | 17682 (0.13) | 42755 (0.1) | 60437 (0.11) |
|  | Severe Depression | 12490 (0.09) | 31337 (0.08) | 43827 (0.08) |
| Drugs | Systemic Corticosteroids | 89295 (0.66) | 207367 (0.51) | 296662 (0.55) |
|  | Antipsychotic Medication | 19455 (0.14) | 43149 (0.11) | 62604 (0.12) |
|  | Benzodiazepine | 43944 (0.33) | 94614 (0.23) | 138558 (0.26) |
| Medical Utilization | Number of Previous Vaccinations: mean (SD) | 1.19 (1.4) | 1.14 (1.37) | 1.15 (1.38) |
|  | Visits per Month: mean (SD) | 1.8 (1.72) | 1.48 (1.66) | 1.56 (1.68) |
|  | 2 Visits Within Year of COVID | 127479 (0.95) | 362941 (0.89) | 490420 (0.9) |
| Socioeconomic Factors | Poverty Rate: mean (SD) | 15.31 (5.19) | 14.91 (5.11) | 15 (5.13) |
|  | Social Deprivation Index Score: mean (SD) | 45.48 (27.15) | 45.1 (28.83) | 45.19 (28.44) |

We analyzed EHR data from 542,938 patients with comorbid depression and COVID-19; 134,595 (74% female) were prescribed SSRIs, and 408,343 (71% female) were not prescribed SSRIs during acute COVID-19. SSRI patients had an average Charlson Comorbidity Index Score of 2.36, while non-SSRI patients had an average of 1.96. Approximately 9% of SSRI patients had severe depression, while 8% of non-SSRI patients had severe depression. Patients prescribed SSRIs had, on average, 1.8 healthcare interactions per month, while those not prescribed SSRIs had 1.48 per month.

We found that patients who were prescribed SSRIs during COVID-19 had a significantly lower risk of gastrointestinal (adjusted risk ratio (aRR) 0.95, 95% CI 0.92, 0.97), general (aRR 0.91, 95% CI 0.88, 0.95), headache (aRR 0.96, 95% CI 0.92, 0.99) reproductive (aRR 0.96, 95% CI 0.93, 1.00), and skin-related (aRR 0.92, 95% CI 0.87, 0.98) symptoms and conditions (Figure 1, Table 2). Reproductive symptoms and conditions did not retain significance after FDR correction. We did not detect a significant relationship between SSRI prescription and cardiovascular (aRR 0.99, 95% CI 0.96, 1.02), ear-related (aRR 0.99, 95% CI 0.95, 1.03), ear-nose-throat-related (aRR 0.96, 95% CI 0.92, 1.01), eye-related (aRR 0.94, 95% CI 0.88, 1.01), autoimmunity-related (aRR 0.98, 95% CI 0.87, 1.11), smell and taste related (aRR 0.95, 95% CI 0.84, 1.07), speech and language-related (aRR 1.04, 95% CI 0.93, 1.16), or pulmonary (aRR, 95% CI 0.97, 1.03) symptoms and conditions. Patients prescribed SSRIs during acute COVID-19 had a significantly higher risk of abnormal laboratory findings (aRR 1.06, 95% CI, 1.12), compared to patients not prescribed an SSRI, but this observation did not retain significance after FDR correction.

**Figure 1.**
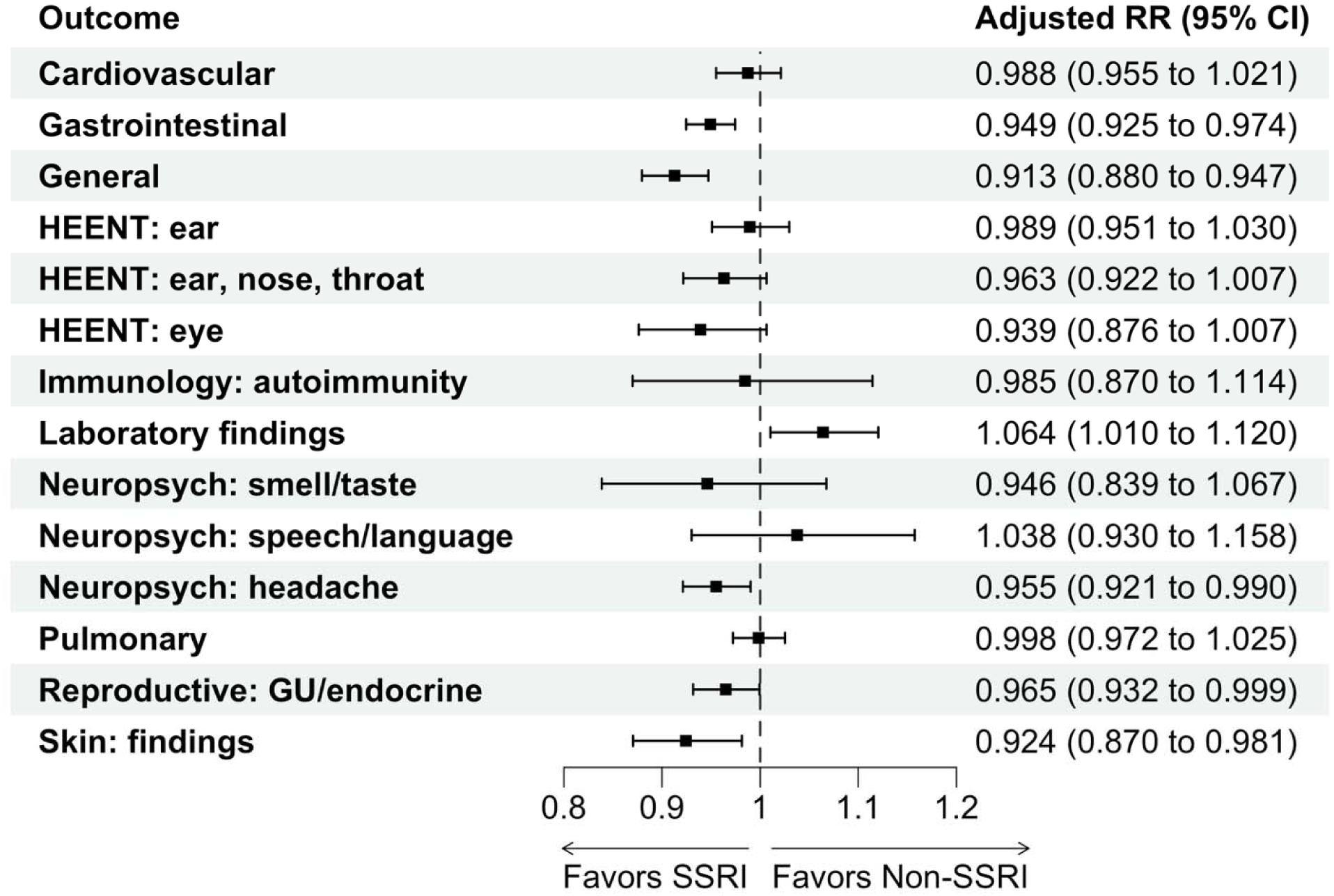
Adjusted relationships between SSRI prescription (*n* = 134,595) during acute COVID-19 and subsequent 12-month cumulative incidence of conditions and symptoms associated with Long COVID, compared to no SSRI prescription (*n* = 408,343), among patients with depression. SSRI: Selective serotonin reuptake inhibitor, RR: Risk ratio, CI: Confidence interval, HEENT: Head, eyes, ears, nose, and throat, Neuropsych: Neuropsychological, GU: genitourinary

**Table 2.** Unadjusted relationships between SSRI prescription (*n* = 134,595) during acute COVID-19 and subsequent 12-month cumulative incidence of conditions and symptoms associated with Long COVID, compared to no SSRI prescription (*n* = 408,343) and false discovery rate corrected p-values, among patients with depression.

| Outcome | Count SSRI Patients | Count Non-SSRI Patients | Number of SSRI Patients with a LC symptom | Number of non-SSRI Patients with a LC symptom | Risk SSRI Patients | Risk Non-SSRI Patients | Unadjusted RR (95% CI) | FDR Corrected Adjusted P value |
| --- | --- | --- | --- | --- | --- | --- | --- | --- |
| Cardiovascular | 134595 | 408343 | 23474 | 55773 | 0.174 | 0.137 | 1.28 (1.26, 1.30) | 0.65 |
| Gastrointestinal (General) | 134595 | 408343 | 39829 | 101047 | 0.296 | 0.247 | 1.20 (1.18, 1.21) | 0 |
| General | 134595 | 408343 | 27308 | 72338 | 0.203 | 0.177 | 1.15 (1.13, 1.16) | 0 |
| HEENT:<br>ear | 134595 | 408343 | 14067 | 33761 | 0.105 | 0.083 | 1.26 (1.24, 1.29) | 0.7 |
| HEENT:<br>ear, nose,<br>and throat | 134595 | 408343 | 8558 | 20725 | 0.064 | 0.051 | 1.25 (1.22, 1.28) | 0.17 |
| HEENT:<br>eye | 134595 | 408343 | 6386 | 16312 | 0.047 | 0.04 | 1.19 (1.15, 1.22) | 0.15 |
| Immunolo<br>gy:<br>autoimmu<br>nity | 134595 | 408343 | 1203 | 2800 | 0.009 | 0.007 | 1.29 (1.22, 1.39) | 0.87 |
| Laborator<br>y findings | 134595 | 408343 | 10887 | 24869 | 0.081 | 0.061 | 1.33 (1.30, 1.36) | 0.05 |
| Neuropsychiatric:<br>smell and<br>taste | 134595 | 408343 | 451 | 996 | 0.003 | 0.002 | 1.37 (1.23, 1.54) | 0.57 |
| Neuropsychiatric:<br>speech<br>and<br>language | 134595 | 408343 | 1295 | 2690 | 0.01 | 0.007 | 1.46 (1.37, 1.56) | 0.65 |
| Neuropsychiatric:<br>headache | 134595 | 408343 | 18012 | 45529 | 0.134 | 0.111 | 1.20 (1.18, 1.22) | 0.04 |
| Pulmonary | 134595 | 408343 | 32218 | 79147 | 0.239 | 0.194 | 1.23 (1.22, 1.25) | 0.91 |
| Reproductive:<br>genitourinary,<br>endocrine,<br>metabolism | 134595 | 408343 | 18746 | 44994 | 0.139 | 0.11 | 1.26 (1.24, 1.28) | 0.11 |
| Skin:<br>findings | 134595 | 408343 | 7574 | 18740 | 0.056 | 0.047 | 1.23 (1.20, 1.26) | 0.04 |

SSRI: Selective serotonin reuptake inhibitor, RR: Risk ratio, CI: Confidence interval, HEENT: Head, eyes, ears, nose, and throat, Neuropsych: Neuropsychological, GU: genitourinary, FDR: false discovery rate

## DISCUSSION

We found that patients who were prescribed SSRIs during acute COVID-19 had a significantly lower risk of subsequent gastrointestinal, headache-related, skin-related, and general symptoms and conditions, compared to patients not prescribed SSRIs, among patients with depression and comorbid COVID-19 between October 1, 2021, and February 1, 2024. These findings are consistent with previous observational studies that have found that the use of SSRIs during acute COVID-19 was associated with a lower risk of Long COVID,^1–3^ although these findings have not been replicated in a randomized context.^4^

These findings support hypotheses about the importance of serotonin, which has key receptors in both the brain and the gut, across several Long COVID phenotypes and pathways.^24^ A key hypothesized mechanism of Long COVID posits that residual SARS-CoV-2 viral particles in Long COVID patients trigger the sustained release of virus-induced Type 1 interferon, which prevents the production of tryptophan, which is a precursor to serotonin.^1,2^ This reduced production of serotonin impairs neurologic and vagal functioning, while contributing to hypercoagulation.^1,2^ *Gastrointestinal* symptoms and conditions are central to this pathway, as serotonin and tryptophan are gut-derived, and vagal functioning is a key component of gut-brain communication.^1,24,25^ Furthermore, impaired vagal functioning and hypercoagulation via insufficient peripheral serotonin may explain the increased incidence of *headaches*, *skin-related*, and *general* symptoms.^24^ Cumulatively, these finding highlights the potential role of serotonin in contributing to disparate Long COVID-related phenotypes.

### Strengths and limitations

Our categorization of patient symptom profiles is a potential limitation of this study. We chose to summarize patient symptom burden across 14 categories as binary indicators (i.e., 14 yes/no values) in order to retain phenotype specificity while maintaining study power and maximizing interpretability. Residual confounding by symptom severity or symptom duration remains plausible, although we lacked reliable data on these metrics. Furthermore, our counterfactual intervention contrasted outcomes between patients with no symptoms versus patients with one or more symptoms, which leaves the possibility of unique relationships among patients with high symptom burden (e.g., 5 or more symptoms), although our sample size of high symptom burden patients was small and therefore evaluations would be vulnerable to near positivity violations. Another limitation is our exclusion of symptoms that were related to depression or SSRI use. Including these symptoms would be vulnerable to measurement bias, as patients prescribed SSRIs would be evaluated for them more frequently. Future studies using active outcome ascertainment (i.e., not vulnerable to measurement bias) should include these symptoms in their assessments. The generalizability of N3C has been described previously as a limitation. While N3C is a broad, national sample, it overrepresents academic research centers and patients with high healthcare-seeking behaviors. Therefore, our sample has a disproportionately high socioeconomic status, is older, and has more comorbidities than the general population.^6,12,26^

The data source, N3C, was a strength of this study, as it includes high-dimensional EHR data from a large patient sample.^8^ The analysis approach, using Super Learner and targeted maximum likelihood estimation, was a second strength of this study.^18,19,22,27^ This approach allows us to flexibly model the outcome regression, treatment mechanism, and censoring mechanism with minimal parametric assumptions while adjusting for confounders of interest. This setting, where we face informative censoring and a difficult-to-characterize treatment mechanism, benefits from the robustness of this approach.

## CONCLUSIONS

We found that the prescription of SSRIs during acute COVID-19 was associated with a significantly lower risk of post-COVID sequelae related to gastrointestinal, headache-related, skin-related, and general symptoms and conditions, compared to no SSRI prescription, among patients with depression and comorbid COVID-19 between October 1, 2021, and February 1, 2024. These findings highlight the role of serotonin in Long COVID and highlight specific sequelae that may be reduced by SSRIs.

## Data Availability

All analytic code is available upon request from the N3C Enclave. Access to study data may be requested in the N3C Enclave as legacy data pending N3C approval. Access to the N3C Data Enclave is managed by NCATS (https://ncats.nih.gov/research/research-activities/n3c/resources/data-access). Interested researchers must first complete a data use agreement and then a data use request to access the N3C Data Enclave. Once access is granted, the N3C data use committee must review and approve all data use, and the publication committee must approve all publications involving N3C data.

## SUPPLEMENTAL MATERIALS

**Supplemental Material 1.** Symptoms and conditions associated with Long COVID.

| HPO_id | HPO_Label | category |
| --- | --- | --- |
| HP:0030784 | Anomic aphasia | neuropsychiatric-speech-language |
| HP:0002381 | Aphasia | neuropsychiatric-speech-language |
| HP:0033849 | Bilingual aphasia | neuropsychiatric-speech-language |
| HP:0002427 | Expressive aphasia | neuropsychiatric-speech-language |
| HP:0033848 | Receptive aphasia | neuropsychiatric-speech-language |
| HP:0001350 | Slurred speech | neuropsychiatric-speech-language |
| HP:0001618 | Dysphonia | HEENT-ENT |
| HP:0001742 | Nasal congestion | HEENT-ENT |
| HP:0033050 | Pharyngalgia | HEENT-ENT |
| HP:0012384 | Rhinitis | HEENT-ENT |
| HP:0010524 | Agnosia | neuropsychiatric-cognitive-dysfunction |
| HP:0002067 | Bradykinesia | neuropsychiatric-cognitive-dysfunction |
| HP:0031843 | Bradyphrenia | neuropsychiatric-cognitive-dysfunction |
| HP:0100543 | Cognitive impairment | neuropsychiatric-cognitive-dysfunction |
| HP:0001289 | Confusion | neuropsychiatric-cognitive-dysfunction |
| HP:0031987 | Diminished ability to concentrate | neuropsychiatric-cognitive-dysfunction |
| HP:0001298 | Encephalopathy | neuropsychiatric-cognitive-dysfunction |
| HP:0033844 | Tachyphrenia | neuropsychiatric-cognitive-dysfunction |
| HP:0001596 | Alopecia | skin-findings |
| HP:0011971 | Dermatographic urticaria | skin-findings |
| HP:0031284 | Flushing | skin-findings |
| HP:0001808 | Fragile nails | skin-findings |
| HP:0000975 | Hyperhidrosis | skin-findings |
| HP:0000967 | Petechiae | skin-findings |
| HP:0000989 | Pruritus | skin-findings |
| HP:0033696 | Pseudo-chilblain | skin-findings |
| HP:0040189 | Scaling skin | skin-findings |
| HP:0000988 | Skin rash | skin-findings |
| HP:0031983 | Abnormal pulmonary thoracic imaging finding | pulmonary-imaging |
| HP:0100750 | Atelectasis | pulmonary-imaging |
| HP:0002110 | Bronchiectasis | pulmonary-imaging |
| HP:0025180 | Centrilobular ground-glass opacification on pulmonary HRCT | pulmonary-imaging |
| HP:0033659 | Crazy-paving pattern | pulmonary-imaging |
| HP:0030879 | Interlobular septal thickening | pulmonary-imaging |
| HP:0032177 | Parenchymal consolidation | pulmonary-imaging |
| HP:0031944 | Pleural thickening | pulmonary-imaging |
| HP:0032446 | Pulmonary bulla | pulmonary-imaging |
| **HP:0002206** | Pulmonary fibrosis | pulmonary-imaging |
| **HP:0033711** | Pulmonary interstitial thickening | pulmonary-imaging |
| **HP:0025390** | Reticular pattern on pulmonary HRCT | pulmonary-imaging |
| **HP:0033609** | Solid pulmonary nodule | pulmonary-imaging |
| **HP:0033610** | Subsolid pulmonary nodule | pulmonary-imaging |
| **HP:0033702** | Subpleural curvilinear line | pulmonary-imaging |
| **HP:0012213** | Decreased glomerular filtration rate | reproductive-genitourinary-endocrine-metabolism |
| **HP:0000819** | Diabetes mellitus | reproductive-genitourinary-endocrine-metabolism |
| **HP:0000969** | Edema | reproductive-genitourinary-endocrine-metabolism |
| **HP:0030014** | Female sexual dysfunction | reproductive-genitourinary-endocrine-metabolism |
| **HP:0001945** | Fever | reproductive-genitourinary-endocrine-metabolism |
| **HP:0002046** | Heat intolerance | reproductive-genitourinary-endocrine-metabolism |
| **HP:0002045** | Hypothermia | reproductive-genitourinary-endocrine-metabolism |
| **HP:0000858** | Irregular menstruation | reproductive-genitourinary-endocrine-metabolism |
| **HP:0011134** | Low-grade fever | reproductive-genitourinary-endocrine-metabolism |
| **HP:0040307** | Male sexual dysfunction | reproductive-genitourinary-endocrine-metabolism |
| **HP:0000132** | Menorrhagia | reproductive-genitourinary-endocrine-metabolism |
| **HP:0001733** | Pancreatitis | reproductive-genitourinary-endocrine-metabolism |
| **HP:0033840** | Postmenopausal bleeding | reproductive-genitourinary-endocrine-metabolism |
| **HP:0001954** | Recurrent fever | reproductive-genitourinary-endocrine-metabolism |
| **HP:0000083** | Renal insufficiency | reproductive-genitourinary-endocrine-metabolism |
| **HP:0005968** | Temperature instability | reproductive-genitourinary-endocrine-metabolism |
| **HP:0033839** | Testicular pain | reproductive-genitourinary-endocrine-metabolism |
| **HP:0000020** | Urinary incontinence | reproductive-genitourinary-endocrine-metabolism |
| **HP:0006536** | Airway obstruction | pulmonary-finding |
| **HP:0045051** | Decreased DLCO | pulmonary-finding |
| **HP:0033760** | Decreased maximal oxygen uptake | pulmonary-finding |
| **HP:0033773** | Decreased RV/TLC ratio | pulmonary-finding |
| **HP:0025179** | Ground-glass opacification | pulmonary-finding |
| **HP:0012418** | Hypoxemia | pulmonary-finding |
| **HP:0030874** | Oxygen desaturation on exertion | pulmonary-finding |
| **HP:0030877** | Reduced FEV1/FVC ratio | pulmonary-finding |
| **HP:0032342** | Reduced forced expiratory volume in one second | pulmonary-finding |
| **HP:0032341** | Reduced forced vital capacity | pulmonary-finding |
| **HP:0033845** | Sense of impending doom | neuropsychiatric-emotion-mood |
| **HP:0031589** | Suicidal ideation | neuropsychiatric-emotion-mood |
| **HP:0033705** | Tearfulness | neuropsychiatric-emotion-mood |
| **HP:0033689** | Anterograde memory impairment | neuropsychiatric-memory |
| **HP:0033688** | Long term memory impairment | neuropsychiatric-memory |
| **HP:0002354** | Memory impairment | neuropsychiatric-memory |
| **HP:0033691** | Procedural memory loss | neuropsychiatric-memory |
| HP:0033687 | Short term memory impairment | neuropsychiatric-memory |
| HP:0002829 | Arthralgia | General-pain |
| HP:0033047 | Body ache | General-pain |
| HP:0002653 | Bone pain | General-pain |
| HP:0100749 | Chest pain | General-pain |
| HP:0033746 | Intrascapular pain | General-pain |
| HP:0009763 | Limb pain | General-pain |
| HP:0003326 | Myalgia | General-pain |
| HP:0033345 | Neuralgia | General-pain |
| HP:0012531 | Pain | General-pain |
| HP:0002020 | Gastroesophageal reflux | gi-findings |
| HP:0002578 | Gastroparesis | gi-findings |
| HP:0002592 | Gastric ulcer | gi-findings |
| HP:0001397 | Hepatic steatosis | gi-findings |
| HP:0002240 | Hepatomegaly | gi-findings |
| HP:0012115 | Hepatitis | gi-findings |
| HP:0004395 | Malnutrition | gi-findings |
| HP:0033757 | Pancreatic steatosis | gi-findings |
| HP:0001744 | Splenomegaly | gi-findings |
| HP:0012053 | Decreased circulating calcifediol concentration | Lab |
| HP:0003155 | Elevated circulating alkaline phosphatase concentration | Lab |
| HP:0031964 | Elevated circulating alanine aminotransferase concentration | Lab |
| HP:0031956 | Elevated circulating aspartate aminotransferase concentration | Lab |
| HP:0003259 | Elevated circulating creatinine concentration | Lab |
| HP:0003236 | Elevated circulating creatine kinase concentration | Lab |
| HP:0011227 | Elevated circulating C-reactive protein concentration | Lab |
| HP:0033106 | Elevated circulating D-dimer concentration | Lab |
| HP:0033833 | Elevated circulating soluble CD25 concentration | Lab |
| HP:0002925 | Elevated circulating thyroid-stimulating hormone concentration | Lab |
| HP:0003565 | Elevated erythrocyte sedimentation rate | Lab |
| HP:0030948 | Elevated gamma-glutamyltransferase level | Lab |
| HP:0002901 | Hypocalcemia | Lab |
| HP:0011900 | Hypofibrinogenemia | Lab |
| HP:0003074 | Hyperglycemia | Lab |
| HP:0001943 | Hypoglycemia | Lab |
| HP:0002148 | Hypophosphatemia | Lab |
| HP:0003281 | Increased circulating ferritin concentration | Lab |
| HP:0030783 | Increased circulating interleukin 6 | Lab |
| HP:0025435 | Increased circulating lactate dehydrogenase concentration | Lab |
| HP:0031185 | Increased circulating NT-proBNP concentration | Lab |
| HP:0032308 | Increased circulating procalcitonin concentration | Lab |
| **HP:0001873** | Thrombocytopenia | Lab |
| **HP:0001681** | Angina pectoris | cardiovascular-symptom |
| **HP:0001962** | Palpitations | cardiovascular-symptom |
| **HP:0001297** | Stroke | cardiovascular-symptom |
| **HP:0001279** | Syncope | cardiovascular-symptom |
| **HP:0000739** | Anxiety | neuropsychiatric-behavioral |
| **HP:0000741** | Apathy | neuropsychiatric-behavioral |
| **HP:0007018** | Attention deficit hyperactivity disorder | neuropsychiatric-behavioral |
| **HP:0008765** | Auditory hallucinations | neuropsychiatric-behavioral |
| **HP:0033630** | Brain fog | neuropsychiatric-behavioral |
| **HP:0000746** | Delusions | neuropsychiatric-behavioral |
| **HP:0000738** | Hallucinations | neuropsychiatric-behavioral |
| **HP:0033051** | Impaired executive functioning | neuropsychiatric-behavioral |
| **HP:0100710** | Impulsivity | neuropsychiatric-behavioral |
| **HP:0000737** | Irritability | neuropsychiatric-behavioral |
| **HP:0025269** | Panic attack | neuropsychiatric-behavioral |
| **HP:0002183** | Phonophobia | neuropsychiatric-behavioral |
| **HP:0001959** | Polydipsia | neuropsychiatric-behavioral |
| **HP:0033676** | Posttraumatic stress symptom | neuropsychiatric-behavioral |
| **HP:0000736** | Short attention span | neuropsychiatric-behavioral |
| **HP:0033694** | Tactile hallucination | neuropsychiatric-behavioral |
| **HP:0002367** | Visual hallucinations | neuropsychiatric-behavioral |
| **HP:0012735** | Cough | pulmonary-symptom |
| **HP:0002094** | Dyspnea | pulmonary-symptom |
| **HP:0033750** | Reduced functional residual capacity | pulmonary-finding |
| **HP:0033753** | Reduced residual volume | pulmonary-finding |
| **HP:0033169** | Reduced total lung capacity | pulmonary-finding |
| **HP:0002091** | Restrictive ventilatory defect | pulmonary-finding |
| **HP:0002102** | Pleuritis | pulmonary-finding |
| **HP:0002204** | Pulmonary embolism | pulmonary-finding |
| **HP:0100845** | Anaphylactic shock | immunology-autoimmunity |
| **HP:0003493** | Antinuclear antibody positivity | immunology-autoimmunity |
| **HP:0025379** | Anti-thyroid peroxidase antibody positivity | immunology-autoimmunity |
| **HP:0032069** | Anti-thyroglobulin antibody positivity | immunology-autoimmunity |
| **HP:0002716** | Lymphadenopathy | immunology-autoimmunity |
| **HP:0001888** | Lymphopenia | immunology-autoimmunity |
| **HP:0002315** | Headache | neuropsychiatric-headache |
| **HP:0002076** | Migraine | neuropsychiatric-headache |
| **HP:0030766** | Ear pain | HEENT-ear |
| **HP:0000365** | Hearing impairment | HEENT-ear |
| **HP:0010780** | Hyperacusis | HEENT-ear |
| **HP:0008629** | Pulsatile tinnitus | HEENT-ear |
| **HP:0000360** | Tinnitus | HEENT-ear |
| **HP:0002321** | Vertigo | HEENT-ear |
| **HP:0041051** | Ageusia | neuropsychiatric-smell-taste |
| **HP:0000458** | Anosmia | neuropsychiatric-smell-taste |
| **HP:0000224** | Hypogeusia | neuropsychiatric-smell-taste |
| **HP:0004409** | Hyposmia | neuropsychiatric-smell-taste |
| **HP:0031249** | Parageusia | neuropsychiatric-smell-taste |
| **HP:0033847** | Phantageusia | neuropsychiatric-smell-taste |
| **HP:0033693** | Phantosmia | neuropsychiatric-smell-taste |
| **HP:0001369** | Arthritis | General-symptom |
| **HP:0025406** | Asthenia | General-symptom |
| **HP:0031352** | Chest tightness | General-symptom |
| **HP:0025143** | Chills | General-symptom |
| **HP:0033850** | Coldness | General-symptom |
| **HP:0002355** | Difficulty walking | General-symptom |
| **HP:0033665** | Diminished health-related quality of life | General-symptom |
| **HP:0033667** | Diminished mental health | General-symptom |
| **HP:0033666** | Diminished physical functioning | General-symptom |
| **HP:0003546** | Exercise intolerance | General-symptom |
| **HP:0012378** | Fatigue | General-symptom |
| **HP:0033675** | Frailty | General-symptom |
| **HP:0031059** | Impaired ability to bathe oneself | General-symptom |
| **HP:0031060** | Impaired ability to dress oneself | General-symptom |
| **HP:0031058** | Impairment of activities of daily living | General-symptom |
| **HP:0033834** | Malaise | General-symptom |
| **HP:0030166** | Night sweats | General-symptom |
| **HP:0033695** | Occupational disability | General-symptom |
| **HP:0030973** | Postexertional malaise | General-symptom |
| **HP:0025144** | Shivering | General-symptom |
| **HP:0025258** | Stiff neck | General-symptom |
| **HP:0001824** | Weight loss | General-symptom |
| **HP:0000217** | Xerostomia | General-symptom |
| **HP:0001662** | Bradycardia | cardiovascular-finding |
| **HP:4000006** | Elevated myocardial native T1 | cardiovascular-finding |
| **HP:4000003** | Elevated myocardial native T2 | cardiovascular-finding |
| **HP:0000822** | Hypertension | cardiovascular-finding |
| **HP:0002615** | Hypotension | cardiovascular-finding |
| **HP:0410174** | Increased circulating troponin T concentration | cardiovascular-finding |
| **HP:0410173** | Increased circulating troponin I concentration | cardiovascular-finding |
| **HP:0031862** | Increased heart rate variability | cardiovascular-finding |
| **HP:0033755** | Increased left ventricular end-diastolic volume | cardiovascular-finding |
| **HP:4000004** | Myocardial late gadolinium enhancement | cardiovascular-finding |
| **HP:0012819** | Myocarditis | cardiovascular-finding |
| **HP:0001698** | Pericardial effusion | cardiovascular-finding |
| **HP:4000005** | Pericardial late gadolinium enhancement | cardiovascular-finding |
| **HP:0012664** | Reduced ejection fraction | cardiovascular-finding |
| **HP:0001649** | Tachycardia | cardiovascular-finding |
| **HP:0004936** | Venous thrombosis | cardiovascular-finding |
| **HP:0000718** | Aggressive behavior | neuropsychiatric-emotion-mood |
| **HP:0000716** | Depression | neuropsychiatric-emotion-mood |
| **HP:0033838** | Dysphoria | neuropsychiatric-emotion-mood |
| **HP:0000712** | Emotional lability | neuropsychiatric-emotion-mood |
| **HP:0031844** | Euphoria | neuropsychiatric-emotion-mood |
| **HP:0100754** | Mania | neuropsychiatric-emotion-mood |
| **HP:0002875** | Exertional dyspnea | pulmonary-symptom |
| **HP:0002105** | Hemoptysis | pulmonary-symptom |
| **HP:0033709** | Increased sputum production | pulmonary-symptom |
| **HP:0031246** | Nonproductive cough | pulmonary-symptom |
| **HP:0033771** | Pleuritic chest pain | pulmonary-symptom |
| **HP:0031245** | Productive cough | pulmonary-symptom |
| **HP:0033710** | Rest dyspnea | pulmonary-symptom |
| **HP:0031417** | Rhinorrhea | pulmonary-symptom |
| **HP:0030831** | Rhonchi | pulmonary-symptom |
| **HP:0025095** | Sneeze | pulmonary-symptom |
| **HP:0002789** | Tachypnea | pulmonary-symptom |
| **HP:0030828** | Wheezing | pulmonary-symptom |
| **HP:0033747** | Abnormal exteroceptive sensation | neuropsychiatric-finding |
| **HP:0031826** | Abnormal reflex | neuropsychiatric-finding |
| **HP:0100022** | Abnormality of movement | neuropsychiatric-finding |
| **HP:0001251** | Ataxia | neuropsychiatric-finding |
| **HP:0003487** | Babinski sign | neuropsychiatric-finding |
| **HP:0001260** | Dysarthria | neuropsychiatric-finding |
| **HP:0001310** | Dysmetria | neuropsychiatric-finding |
| **HP:0002015** | Dysphagia | neuropsychiatric-finding |
| **HP:0001332** | Dystonia | neuropsychiatric-finding |
| **HP:0007209** | Facial paralysis | neuropsychiatric-finding |
| **HP:0000743** | Frontal release signs | neuropsychiatric-finding |
| **HP:0001288** | Gait disturbance | neuropsychiatric-finding |
| **HP:0030237** | Hand muscle weakness | neuropsychiatric-finding |
| **HP:0100963** | Hyperesthesia | neuropsychiatric-finding |
| **HP:0002487** | Hyperkinetic movements | neuropsychiatric-finding |
| **HP:0033748** | Hypoesthesia | neuropsychiatric-finding |
| **HP:0001252** | Hypotonia | neuropsychiatric-finding |
| **HP:0003394** | Muscle spasm | neuropsychiatric-finding |
| **HP:0001324** | Muscle weakness | neuropsychiatric-finding |
| **HP:0001278** | Orthostatic hypotension | neuropsychiatric-finding |
| **HP:0003401** | Paresthesia | neuropsychiatric-finding |
| **HP:0001300** | Parkinsonism | neuropsychiatric-finding |
| **HP:0001271** | Polyneuropathy | neuropsychiatric-finding |
| **HP:0002063** | Rigidity | neuropsychiatric-finding |
| **HP:0001250** | Seizure | neuropsychiatric-finding |
| **HP:0003202** | Skeletal muscle atrophy | neuropsychiatric-finding |
| **HP:0003474** | Somatic sensory dysfunction | neuropsychiatric-finding |
| **HP:0001257** | Spasticity | neuropsychiatric-finding |
| **HP:0001337** | Tremor | neuropsychiatric-finding |
| **HP:0012799** | Unilateral facial palsy | neuropsychiatric-finding |
| **HP:0100785** | Insomnia | neuropsychiatric-sleep |
| **HP:0031355** | Maintenance insomnia | neuropsychiatric-sleep |
| **HP:0012452** | Restless legs | neuropsychiatric-sleep |
| **HP:0010535** | Sleep apnea | neuropsychiatric-sleep |
| **HP:0002360** | Sleep disturbance | neuropsychiatric-sleep |
| **HP:0031354** | Sleep onset insomnia | neuropsychiatric-sleep |
| **HP:0031356** | Terminal insomnia | neuropsychiatric-sleep |
| **HP:0002027** | Abdominal pain | gi-symptoms |
| **HP:0011458** | Abdominal symptom | gi-symptoms |
| **HP:0002039** | Anorexia | gi-symptoms |
| **HP:0002607** | Bowel incontinence | gi-symptoms |
| **HP:0002019** | Constipation | gi-symptoms |
| **HP:0002014** | Diarrhea | gi-symptoms |
| **HP:0033842** | Early satiety | gi-symptoms |
| **HP:0002018** | Nausea | gi-symptoms |
| **HP:0002013** | Vomiting | gi-symptoms |
| **HP:0004396** | Poor appetite | gi-symptoms |
| **HP:0000618** | Blindness | HEENT-eye |
| **HP:0000622** | Blurred vision | HEENT-eye |
| **HP:0000509** | Conjunctivitis | HEENT-eye |
| **HP:0000651** | Diplopia | HEENT-eye |
| **HP:0000640** | Gaze-evoked nystagmus | HEENT-eye |
| **HP:0001097** | Keratoconjunctivitis sicca | HEENT-eye |
| **HP:0200026** | Ocular pain | HEENT-eye |
| **HP:0033841** | Ocular pruritus | HEENT-eye |
| **HP:0007994** | Peripheral visual field loss | HEENT-eye |
| **HP:0000613** | Photophobia | HEENT-eye |
| **HP:0025337** | Red eye | HEENT-eye |
| **HP:0000572** | Visual loss | HEENT-eye |
| **HP:0100832** | Vitreous floaters | HEENT-eye |

## DECLARATIONS

## Ethics approval and consent to participate

This study was approved by the UC Berkeley Office for Protection of Human Subjects (2022-01-14980). The N3C data transfer to NCATS is performed under a Johns Hopkins University Reliance Protocol # IRB00249128 or individual site agreements with NIH. N3C received a waiver of consent from the NIH Institutional Review board and allows the secondary analysis of these data without additional consent.

## Consent to publish

The authors consent to the publication of this manuscript in its entirety.

## Availability of data and materials

All analytic code is available upon request from the N3C Enclave. Access to study data may be requested in the N3C Enclave as “legacy data” pending N3C approval. Access to the N3C Data Enclave is managed by NCATS (https://ncats.nih.gov/research/research-activities/n3c/resources/data-access). Interested researchers must first complete a data use agreement and then a data use request to access the N3C Data Enclave. Once access is granted, the N3C data use committee must review and approve all data use, and the publication committee must approve all publications involving N3C data.

## Authors statement

Authorship was determined using ICMJE recommendations.

ZB: Generated research question, drafted manuscript, managed project timeline, and coordinated analysis.

YJ, LW, MK, JA, EH, AB, RCP, AH, and JH: Provided oversight on study design and analysis plan, supported analysis, reviewed manuscript, and provided feedback.

## Inclusion and ethics statement

All co-authors and collaborators included in this manuscript have fulfilled the criteria for authorship.

## N3C Attribution

The analyses described in this manuscript were conducted with data or tools accessed through the NCATS N3C Data Enclave (RP-884840) https://covid.cd2h.org and N3C Attribution & Publication Policy v 1.2-2020-08-25b supported by NCATS U24 TR002306, Axle Informatics Subcontract: NCATS-P00438-B, the Bill & Melinda Gates Foundation: OPP1165144, and the National Institutes of General Medical Sciences: U54GM115458 and 5U54GM104942-04. This research was possible because of the patients whose information is included within the data and the organizations (https://ncats.nih.gov/n3c/resources/data-contribution/data-transfer-agreement-signatories) and scientists who have contributed to the ongoing development of this community resource [https://doi.org/10.1093/jamia/ocaa196].

## Disclaimer

The N3C Publication Committee confirmed that this manuscript (MSID 2770.413) complies with N3C data use and attribution policies; however, the authors are solely responsible for its content, which does not necessarily represent the official views of the National Institutes of Health or the N3C program.

## IRB

The N3C data transfer to NCATS is performed under a Johns Hopkins University Reliance Protocol # IRB00249128 or individual site agreements with NIH. The N3C Data Enclave is managed under the authority of the NIH; information can be found at https://ncats.nih.gov/n3c/resources.

This research project was approved by the University of California, Berkeley Committee for the Protection of Human Subjects (CPHS protocol number 2022-01-14980). This approval is issued under University of California, Berkeley Federalwide Assurance #00006252.

## Individual Acknowledgements For Core Contributors

We gratefully acknowledge the following core contributors to N3C: Adam B. Wilcox, Adam M. Lee, Alexis Graves, Alfred (Jerrod) Anzalone, Amin Manna, Amit Saha, Amy Olex, Andrea Zhou, Andrew E. Williams, Andrew Southerland, Andrew T. Girvin, Anita Walden, Anjali A. Sharathkumar, Benjamin Amor, Benjamin Bates, Brian Hendricks, Brijesh Patel, Caleb Alexander, Carolyn Bramante, Cavin Ward-Caviness, Charisse Madlock-Brown, Christine Suver, Christopher Chute, Christopher Dillon, Chunlei Wu, Clare Schmitt, Cliff Takemoto, Dan Housman, Davera Gabriel, David A. Eichmann, Diego Mazzotti, Don Brown, Eilis Boudreau, Elaine Hill, Elizabeth Zampino, Emily Carlson Marti, Emily R. Pfaff, Evan French, Farrukh M Koraishy, Federico Mariona, Fred Prior, George Sokos, Greg Martin, Harold Lehmann, Heidi Spratt, Hemalkumar Mehta, Hongfang Liu, Hythem Sidky, J.W. Awori Hayanga, Jami Pincavitch, Jaylyn Clark, Jeremy Richard Harper, Jessica Islam, Jin Ge, Joel Gagnier, Joel H. Saltz, Joel Saltz, Johanna Loomba, John Buse, Jomol Mathew, Joni L. Rutter, Julie A. McMurry, Justin Guinney, Justin Starren, Karen Crowley, Katie Rebecca Bradwell, Kellie M. Walters, Ken Wilkins, Kenneth R. Gersing, Kenrick Dwain Cato, Kimberly Murray, Kristin Kostka, Lavance Northington, Lee Allan Pyles, Leonie Misquitta, Lesley Cottrell, Lili Portilla, Mariam Deacy, Mark M. Bissell, Marshall Clark, Mary Emmett, Mary Morrison Saltz, Matvey B. Palchuk, Melissa A. Haendel, Meredith Adams, Meredith Temple-O’Connor, Michael G. Kurilla, Michele Morris, Nabeel Qureshi, Nasia Safdar, Nicole Garbarini, Noha Sharafeldin, Ofer Sadan, Patricia A. Francis, Penny Wung Burgoon, Peter Robinson, Philip R.O. Payne, Rafael Fuentes, Randeep Jawa, Rebecca Erwin-Cohen, Rena Patel, Richard A. Moffitt, Richard L. Zhu, Rishi Kamaleswaran, Robert Hurley, Robert T. Miller, Saiju Pyarajan, Sam G. Michael, Samuel Bozzette, Sandeep Mallipattu, Satyanarayana Vedula, Scott Chapman, Shawn T. O’Neil, Soko Setoguchi, Stephanie S. Hong, Steve Johnson, Tellen D. Bennett, Tiffany Callahan, Umit Topaloglu, Usman Sheikh, Valery Gordon, Vignesh Subbian, Warren A. Kibbe, Wenndy Hernandez, Will Beasley, Will Cooper, William Hillegass, Xiaohan Tanner Zhang. Details of contributions available at covid.cd2h.org/core-contributors

## Data Partners with Released Data

The following institutions whose data is released or pending:

Available: Advocate Health Care Network — UL1TR002389: The Institute for Translational Medicine (ITM) • Boston University Medical Campus — UL1TR001430: Boston University Clinical and Translational Science Institute • Brown University — U54GM115677: Advance Clinical Translational Research (Advance-CTR) • Carilion Clinic — UL1TR003015: iTHRIV Integrated Translational health Research Institute of Virginia • Charleston Area Medical Center — U54GM104942: West Virginia Clinical and Translational Science Institute (WVCTSI) • Children’s Hospital Colorado — UL1TR002535: Colorado Clinical and Translational Sciences Institute • Columbia University Irving Medical Center — UL1TR001873: Irving Institute for Clinical and Translational Research • Duke University — UL1TR002553: Duke Clinical and Translational Science Institute • George Washington Children’s Research Institute — UL1TR001876: Clinical and Translational Science Institute at Children’s National (CTSA-CN) • George Washington University — UL1TR001876: Clinical and Translational Science Institute at Children’s National (CTSA-CN) • Indiana University School of Medicine — UL1TR002529: Indiana Clinical and Translational Science Institute • Johns Hopkins University — UL1TR003098: Johns Hopkins Institute for Clinical and Translational Research • Loyola Medicine — Loyola University Medical Center • Loyola University Medical Center — UL1TR002389: The Institute for Translational Medicine (ITM) • Maine Medical Center — U54GM115516: Northern New England Clinical & Translational Research (NNE-CTR) Network • Massachusetts General Brigham — UL1TR002541: Harvard Catalyst • Mayo Clinic Rochester — UL1TR002377: Mayo Clinic Center for Clinical and Translational Science (CCaTS) • Medical University of South Carolina — UL1TR001450: South Carolina Clinical & Translational Research Institute (SCTR) • Montefiore Medical Center — UL1TR002556: Institute for Clinical and Translational Research at Einstein and Montefiore • Nemours — U54GM104941: Delaware CTR ACCEL Program • NorthShore University HealthSystem — UL1TR002389: The Institute for Translational Medicine (ITM) • Northwestern University at Chicago — UL1TR001422: Northwestern University Clinical and Translational Science Institute (NUCATS) • OCHIN — INV-018455: Bill and Melinda Gates Foundation grant to Sage Bionetworks • Oregon Health & Science University — UL1TR002369: Oregon Clinical and Translational Research Institute • Penn State Health Milton S. Hershey Medical Center — UL1TR002014: Penn State Clinical and Translational Science Institute • Rush University Medical Center — UL1TR002389: The Institute for Translational Medicine (ITM) • Rutgers, The State University of New Jersey — UL1TR003017: New Jersey Alliance for Clinical and Translational Science • Stony Brook University — U24TR002306 • The Ohio State University — UL1TR002733: Center for Clinical and Translational Science • The State University of New York at Buffalo — UL1TR001412: Clinical and Translational Science Institute • The University of Chicago — UL1TR002389: The Institute for Translational Medicine (ITM) • The University of Iowa — UL1TR002537: Institute for Clinical and Translational Science • The University of Miami Leonard M. Miller School of Medicine — UL1TR002736: University of Miami Clinical and Translational Science Institute • The University of Michigan at Ann Arbor — UL1TR002240: Michigan Institute for Clinical and Health Research • The University of Texas Health Science Center at Houston — UL1TR003167: Center for Clinical and Translational Sciences (CCTS) • The University of Texas Medical Branch at Galveston — UL1TR001439: The Institute for Translational Sciences • The University of Utah — UL1TR002538: Uhealth Center for Clinical and Translational Science • Tufts Medical Center — UL1TR002544: Tufts Clinical and Translational Science Institute • Tulane University — UL1TR003096: Center for Clinical and Translational Science • University Medical Center New Orleans — U54GM104940: Louisiana Clinical and Translational Science (LA CaTS) Center • University of Alabama at Birmingham — UL1TR003096: Center for Clinical and Translational Science • University of Arkansas for Medical Sciences — UL1TR003107: UAMS Translational Research Institute • University of Cincinnati — UL1TR001425: Center for Clinical and Translational Science and Training • University of Colorado Denver, Anschutz Medical Campus — UL1TR002535: Colorado Clinical and Translational Sciences Institute • University of Illinois at Chicago — UL1TR002003: UIC Center for Clinical and Translational Science • University of Kansas Medical Center — UL1TR002366: Frontiers: University of Kansas Clinical and Translational Science Institute • University of Kentucky — UL1TR001998: UK Center for Clinical and Translational Science • University of Massachusetts Medical School Worcester — UL1TR001453: The UMass Center for Clinical and Translational Science (UMCCTS) • University of Minnesota — UL1TR002494: Clinical and Translational Science Institute • University of Mississippi Medical Center — U54GM115428: Mississippi Center for Clinical and Translational Research (CCTR) • University of Nebraska Medical Center — U54GM115458: Great Plains IDeA-Clinical & Translational Research • University of North Carolina at Chapel Hill — UL1TR002489: North Carolina Translational and Clinical Science Institute • University of Oklahoma Health Sciences Center — U54GM104938: Oklahoma Clinical and Translational Science Institute (OCTSI) • University of Rochester — UL1TR002001: UR Clinical & Translational Science Institute • University of Southern California — UL1TR001855: The Southern California Clinical and Translational Science Institute (SC CTSI) • University of Vermont — U54GM115516: Northern New England Clinical & Translational Research (NNE-CTR) Network • University of Virginia — UL1TR003015: iTHRIV Integrated Translational health Research Institute of Virginia • University of Washington — UL1TR002319: Institute of Translational Health Sciences • University of Wisconsin-Madison — UL1TR002373: UW Institute for Clinical and Translational Research • Vanderbilt University Medical Center — UL1TR002243: Vanderbilt Institute for Clinical and Translational Research • Virginia Commonwealth University — UL1TR002649: C. Kenneth and Dianne Wright Center for Clinical and Translational Research • Wake Forest University Health Sciences — UL1TR001420: Wake Forest Clinical and Translational Science Institute • Washington University in St. Louis — UL1TR002345: Institute of Clinical and Translational Sciences • Weill Medical College of Cornell University — UL1TR002384: Weill Cornell Medicine Clinical and Translational Science Center • West Virginia University — U54GM104942: West Virginia Clinical and Translational Science Institute (WVCTSI)

Submitted: Icahn School of Medicine at Mount Sinai — UL1TR001433: ConduITS Institute for Translational Sciences • The University of Texas Health Science Center at Tyler — UL1TR003167: Center for Clinical and Translational Sciences (CCTS) • University of California, Davis — UL1TR001860: UCDavis Health Clinical and Translational Science Center • University of California, Irvine — UL1TR001414: The UC Irvine Institute for Clinical and Translational Science (ICTS) • University of California, Los Angeles — UL1TR001881: UCLA Clinical Translational Science Institute • University of California, San Diego — UL1TR001442: Altman Clinical and Translational Research Institute • University of California, San Francisco — UL1TR001872: UCSF Clinical and Translational Science Institute

Pending: Arkansas Children’s Hospital — UL1TR003107: UAMS Translational Research Institute • Baylor College of Medicine — None (Voluntary) • Children’s Hospital of Philadelphia — UL1TR001878: Institute for Translational Medicine and Therapeutics • Cincinnati Children’s Hospital Medical Center — UL1TR001425: Center for Clinical and Translational Science and Training • Emory University — UL1TR002378: Georgia Clinical and Translational Science Alliance • HonorHealth — None (Voluntary) • Loyola University Chicago — UL1TR002389: The Institute for Translational Medicine (ITM) • Medical College of Wisconsin — UL1TR001436: Clinical and Translational Science Institute of Southeast Wisconsin • MedStar Health Research Institute — UL1TR001409: The Georgetown-Howard Universities Center for Clinical and Translational Science (GHUCCTS) • MetroHealth — None (Voluntary) • Montana State University — U54GM115371: American Indian/Alaska Native CTR • NYU Langone Medical Center — UL1TR001445: Langone Health’s Clinical and Translational Science Institute • Ochsner Medical Center — U54GM104940: Louisiana Clinical and Translational Science (LA CaTS) Center • Regenstrief Institute — UL1TR002529: Indiana Clinical and Translational Science Institute • Sanford Research — None (Voluntary) • Stanford University — UL1TR003142: Spectrum: The Stanford Center for Clinical and Translational Research and Education • The Rockefeller University — UL1TR001866: Center for Clinical and Translational Science • The Scripps Research Institute — UL1TR002550: Scripps Research Translational Institute • University of Florida — UL1TR001427: UF Clinical and Translational Science Institute • University of New Mexico Health Sciences Center — UL1TR001449: University of New Mexico Clinical and Translational Science Center • University of Texas Health Science Center at San Antonio — UL1TR002645: Institute for Integration of Medicine and Science • Yale New Haven Hospital — UL1TR001863: Yale Center for Clinical Investigation

